# Changes in the Genital Microbiome and Inflammation Among Women Using 30-Day *L. Rhamnosus/L. Reuteri* Probiotic in Combination with Intravaginal Estradiol: Findings from a Pilot, Open-Label, Phase I Clinical Trial

**DOI:** 10.64898/2025.12.03.25341486

**Authors:** Smritee Dabee, Maysa Niazy, Biban Gill, Jocelyn M. Wessels, Christina L. Hayes, Aisha Nazli, Jenna Ratcliffe, Junic Wokuri, Gregor Reid, Rupert Kaul, Jesleen Rana, Muna Alkhaifi, Wangari Tharao, Fiona Smaill, Kaushic Charu

## Abstract

A dysbiotic genital microbiome can cause inflammation, higher sexually transmitted infection risk, and long-term effects even when asymptomatic. Probiotic *Lactobacillus* species can potentially improve the genital microbial environment, especially among women with bacterial vaginosis (BV). Further, the hormone estradiol has been linked to mucosal health and better *Lactobacillus* colonization. We evaluated the impact of intravaginal estradiol, with/without use of *Lacticaseibacillus rhamnosus* GR-1 and *Limosilactobacillus reuteri* RC-14, administered vaginally or orally, on the genital microbiome and inflammation.

Through this pilot Phase I, randomized, open-label trial, fifty premenopausal African, Caribbean, and Black women, were randomized to four study arms: intravaginal estradiol (Estring©; 7.5mg/day); vaginal probiotic (RepHresh Pro-B™) twice daily; combination of Estring© and vaginal RepHresh Pro-B™ (twice daily); or the Estring© and oral RepHresh Pro-B™ (twice daily), for 30 days. Baseline BV status was determined by Nugent scoring. The genital microbial composition was measured by 16S rRNA gene sequencing, and inflammation using a 48-plex Luminex cytokine assay.

By mid-intervention, probiotic *Lactobacillus* taxa were detected in the genital tract in the vaginal probiotic only (p<0.01) and vaginal probiotic + Estring arms (p=0.03), but not with oral administration, although abundances returned to baseline levels post-treatment. 51.3% of participants experienced an overall positive shift in microbial composition by end-of-treatment. At follow-up, despite the decrease in probiotic species abundances, total *Lactobacillus* abundance, mostly *L. iners*, remained higher compared to pre-treatment (68.9% vs 60.9%), with lower overall alpha diversity (p=0.04). Further, this change was accompanied by a consistent decrease in inflammation from baseline to end-of-intervention (p=0.04), and 7 days post-treatment (p=0.05), significantly so at all visits among those with an overall positive microbial shift (all p<0.05).

Overall, our findings demonstrate that microbiome-targeted interventions have the capacity to positively modulate the genital microbial and inflammatory milieu, with factors such as the route of administration playing a key role.

**Panel: Research in context:** *Evidence before this study:* The vaginal microbiome is a critical determinant of reproductive health and HIV susceptibility. Bacterial vaginosis (BV) is a genital condition characterized by the depletion of protective *Lactobacillus* species and an overabundance of dysbiotic anaerobes, which leads to high inflammation levels and increased risk of sexually transmitted infection acquisition. Treating with antibiotics, which is the standard of care for BV, leads to recurrence rates exceeding 50% within a year. Promising studies assessing the effectiveness of live probiotics have been shown in some studies to decrease BV recurrence rates, but have yielded inconsistent results, especially with oral delivery. Additionally, low-dose intravaginal estrogen was found to enhance *Lactobacillus* dominance in postmenopausal women. However, its combined effect with probiotics in premenopausal women, including among African, Caribbean, and Black (ACB) women who are disproportionately affected by BV and HIV, remains poorly understood.

*Added value of this study:* This randomized, open-label, phase I trial was, to our knowledge, the first to compare oral versus vaginal delivery of *L. rhamnosus* GR-1 and *L. reuteri* RC-14, with or without intravaginal estrogen, among premenopausal ACB women. Vaginally-administered probiotics were successfully detected in the vaginal tract, albeit temporarily, led to increased *Lactobacillus* abundance, reduced BV-associated anaerobes abundance, and decreased overall genital inflammation levels. Importantly, these shifts in microbial composition, including an increase in *Lactobacillus* spp. abundance persisted beyond treatment with vaginal probiotic use.

*Implications of all the available evidence:* Our findings suggest that vaginal probiotic use with/without estrogen can support the establishment of a more optimal vaginal microbiome and reduce inflammation - two factors central to genital health and HIV prevention. Route of administration was key to detection of the probiotic taxa in the genital tract. Larger studies are needed to determine the longer-term impact of these microbial and immune improvements on BV recurrence and risk of sexually transmitted infection acquisition.

## Introduction

The vaginal microbiota is a key determinant of reproductive and sexual health. A more optimal microbiota is characterized by the dominance of *Lactobacillus* species and low overall diversity, which maintains a protective acidic pH and inhibits pathogenic bacterial growth^1,2^. In contrast, vaginal microbial dysbiosis, particularly bacterial vaginosis (BV), is marked by a depletion of lactobacilli, an overgrowth of pathogenic anaerobic bacteria, and high microbial diversity^3,4^. BV affects nearly 23-29% of reproductive-aged women globally^5^. Further, BV is a well-established risk factor for sexually transmitted infection (STI) acquisition^6^, likely due to damage caused by persistent inflammation, mucosal barrier disruption, decreases in lactic acid concentrations, and several other factors^7^. BV has been shown to increase the risk of HIV acquisition by approximately 60%^8^, primarily caused by *Lactobacillus* spp. depletion^9^. This burden remains heavily borne by women and adolescent girls, who accounted for 44% of all new infections in 2023^10^.

African, Caribbean, and Black (ACB) women are often more likely to have genital microbiomes colonized by BV-associated bacteria and fewer protective lactobacilli, with disproportionately higher prevalences of BV reaching 50%^7,11–13^. In an earlier key study, about 40% of Black and Hispanic women had a highly diverse microbiome dominated by bacteria other than lactobacilli compared to ∼10% among White women and ∼20% among women of Asian ethnicity^14^. Although the exact mechanism remains unclear for these differences, a range of socio-behavioural factors and lived experiences, along with genetics, could be playing a role^15–17^. Even when asymptomatic, microbial dysbiosis can lead to persistent inflammation, with elevated levels of pro-inflammatory cytokines, including IL-1α, IL-1β, TNF-α, IL-6, and IL-8, and long-term sequelae^12,13,18^. Finding ways to prevent this dysbiosis would not only improve gynecological health but would also potentially lower STI and HIV acquisition risk at a population level.

There is no definitive cure for the microbial dysbiosis associated with BV, and first-line treatments, including oral and intravaginal antibiotics, have been associated with recurrence rates exceeding 50% within a year^19^. Hence, alternative strategies that can lead to sustained colonization by *Lactobacillus*-dominated communities are critically needed. Two promising approaches are hormonal therapy and the use of live *Lactobacillus*-based probiotics. Low-dose intravaginal estrogen led to improved epithelial health and glycogen production, thereby supporting *Lactobacillus* dominance, particularly in postmenopausal women^20^. The use of probiotic capsules containing *Lacticaseibacillus rhamnosus* GR-1 and *Limosilactobacillus reuteri* RC-14, alongside antibiotics, has been linked to improved BV cure rates^21,22^. Other trials have explored *Lactobacillus*-based probiotic supplementation for clearance of high-risk human papillomavirus^23^ and enhancement of fertility outcomes^24,25^, indicating that probiotic treatment can have benefits that extend beyond BV clearance. In a landmark study, vaginally-administered probiotic *L. crispatus* use post metronidazole treatment led to a 34% decreased risk of recurrent BV over 12 weeks^26^. However, overall outcomes remain inconsistent across studies exploring different routes of treatment administration, with oral probiotics rarely achieving reliable genital colonization^27,28^. Further, no studies have examined the effectiveness of combining estrogen and probiotics to improve the vaginal health of women.

Currently, there is insufficient data describing how probiotic and hormonal interventions shape both the vaginal microbiome and mucosal immune milieu in premenopausal women. To address this gap, we conducted a randomized, open-label, phase I trial, with the primary aim of assessing the feasibility, safety, and tolerability of administering a low-dose estradiol vaginal ring, administered alone or in combination with *L. rhamnosus* GR-1 and *L. reuteri* RC-14, delivered orally or vaginally^29^. Here, in a secondary analysis, we determine the influence of these interventions on the genital microbiome composition and its dynamic relationship with genital inflammation. We compare oral versus vaginal probiotic delivery, in combination with estrogen, on both microbial and immune outcomes, among ACB women, a population with disproportionately higher rates of vaginal microbial dysbiosis and higher prevalence of HIV.

## Methods

### Study design and participant recruitment

As part of the parent study, previously described by Gill et al. (2025)^30^, fifty African, Caribbean, and Black (ACB) women from the Greater Toronto Area were recruited between November 2019 and December 2021. Here, in the absence of gender identity information, the term women was used to describe individuals with a cervix. Briefly, participants attended five clinic visits at the Women’s Health in Women’s Hands (WHIWH) Community Centre in Ontario, Canada, as part of a 30-day, prospective, randomized, open-label, phase I trial with a 7-day follow-up. They were eligible if they were aged 18–49 years old, premenopausal, in good general health, and HIV-negative. They could not have been using an oral probiotic, antibiotic, or steroids in the last 30 days, could not be pregnant, be on hormonal contraception for the past three months, or be using an intra-uterine device, test positive for *Neisseria gonorrhoea* and/or *Chlamydia trachomatis,* or be symptomatic for other STIs or vulvovaginal candidiasis. Participants were instructed to refrain from vaginal intercourse for 48 hours before attending clinic visits. Detailed inclusion and exclusion criteria are described in the parent manuscript^30^. Ethics approval was obtained from the Hamilton Integrated Research Ethics Board (HiREB Project #7061), and the parent trial was registered with Clinicaltrials.gov (NCT03837015) and CIHR Canadian HIV Clinical Trials Network (CTN 308).

To evaluate the feasibility, safety, and tolerability of low-dose intravaginal estrogen and/or *Lactobacillus* probiotics, participants were randomized in a 1:1:1:1 ratio via a computer-generated block randomization scheme, to either (A) 30-day use of a low-dose intravaginal estrogen-containing vaginal rings (Estring©), (B) Estring in combination with a *Lacticaseibacillus rhamnosus* GR-1 and *Limosilactobacillus reuteri* RC-14 probiotic (RepHresh™ Pro-B™) administered twice daily either orally, (C) Estring in combination with the probiotic administered vaginally, and (D) Vaginal probiotic alone (flow diagram in Figure 1). The visits included a Screening visit (visit 1), a Baseline visit at which the products were first administered (visit 2), a mid-intervention visit (visit 3), a 30-day end-of-intervention visit (visit 4), and a safety visit 7 days post-intervention (visit 5). Data from the screening visit was not included in this analysis.

**Fig 1.**
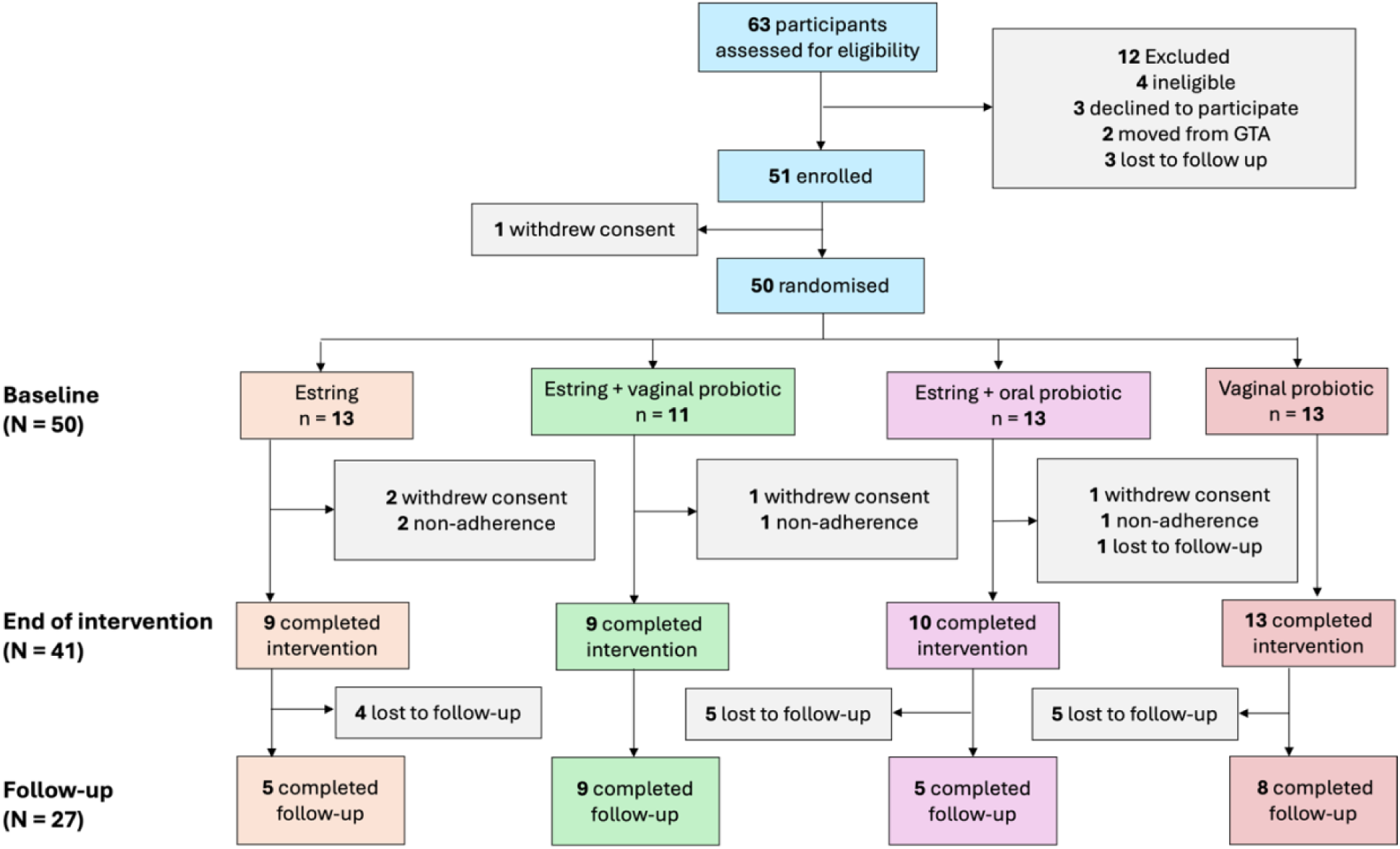
Flow diagram of the trial profile, including enrolment, randomisation, and follow-up of participants included in the intervention cohort.

### Sample collection

At the baseline visit, participants completed a questionnaire covering demographics and sexual history, provided vaginal swabs and a cervicovaginal lavage (CVL) for bacterial vaginosis diagnosis by Nugent scoring,16S rRNA microbiome sequencing, and for measurement of soluble markers (cytokines) by Luminex, as described below.

At baseline, using a speculum, clinician-collected vaginal swabs were obtained by rotating a sterile nylon-flocked swab (Fisher Scientific, CAT# 23-600-960) against the four quadrants of the posterior fornix and across the cervix. A cervicovaginal lavage (CVL) sample was collected by rinsing the endocervix with 2 mL of sterile phosphate-buffered saline (PBS) four times, then aspirating fluid from the posterior fornix. The lavage was centrifuged, and the supernatant was aliquoted into sterile cryovials. All samples were maintained at 4°C and transported to McMaster University for processing.

### Measuring BV status and presence of prostate-specific antigen

At the baseline visit, two additional vaginal swabs were collected, one for BV diagnosis by wet mount and the other to measure prostate-specific antigen (PSA), as a proxy of recent sexual intercourse. BV was diagnosed by Nugent scoring where individuals were given a score of 0-3 for normal vaginal flora, 4-6 for intermediate flora, and 7-10 for BV. To measure PSA levels, swab tips were vortexed 15 times in 0.5 mL of buffer, centrifuged, and tested using the PSA Semiquant test (SERATEC Gesellschaft für Biotechnologie mbH), following the manufacturer’s protocol. Values were reported as presence or absence of PSA.

### Vaginal microbiota and cervicovaginal lavage (CVL) sample collection

Vaginal swabs were processed by cutting the swab tips into sterile 1.5 mL Eppendorf tubes using sterile scissors. Swabs were pulse vortexed 15 times in 0.5 mL sterile PBS, and this was repeated to obtain a total volume of 1.0 mL. The resulting swab tip and fluid were stored at −80°C until DNA extraction.

### Microbial DNA extraction and 16S rRNA gene sequencing

Genomic DNA was extracted from CVL samples using a previously validated protocol designed for low-biomass specimens^31^. 0.1 mm glass beads, GES solution, and monobasic sodium phosphate buffer were added to screw-cap tubes along with 1 mL of the thawed CVL fluid. Mechanical disruption was conducted using a bead beater at 3000 rpm for 3 minutes. Samples then underwent sequential enzymatic digestion, beginning with lysozyme and RNase A at 37°C for 1 hour, followed by proteinase K, SDS, and NaCl at 65°C for 1 hour. Supernatants were collected post-centrifugation and subjected to organic extraction with phenol:chloroform:isoamyl alcohol. The aqueous phase was recovered and purified using the Zymo DNA Clean and Concentrator-25 Kit (Cedarlane, CAT# D4034), and the DNA was eluted into ultrapure water for downstream microbiota analysis.

A nested PCR approach was used to amplify the V3-V4 region of the bacterial 16S rRNA gene, as previously described^31^. Briefly, 200 ng of purified genomic DNA was used for the primary PCR reaction using the primers: 8f (AGAGTTTGATCCTGGCTCAG) and 926r (CCGTCAATTCCTTTRAGTTT). PCR reactions were run in triplicate. Triplicate reactions were pooled, and 3 μL of the combined product was used as a template for the indexing PCR step targeting the V3-V4 region with barcoded Illumina-adapted primers 341f (CCTACGGGNGGCWGCAG) and 806r (GGACTACHVGGGTWTCTAAT)^32^. PCR amplicons were resolved on a 1.5% agarose gel to confirm product size, then normalized using the SequalPrep™ Normalization Kit (ThermoFisher, CAT# A1051001) as per the manufacturer’s guidelines. Sequencing was performed on the Illumina MiSeq instrument using V3 600-cycle kits.

Following demultiplexing, raw V3-V4 region reads were processed by trimming adapter and primer sequences using Cutadapt v5.1^33^. Trimmed reads were then processed with DADA2 v1.16^34^ to infer amplicon sequence variants (ASVs), and a threshold cutoff of 1000 reads was applied. Initial taxonomic classification of ASVs was performed using the SILVA reference database (release 138.1)^35^, with further annotation using BLAST for better species-level resolution. For downstream analyses, the dataset was filtered to retain ASVs with more than five reads in at least 10% of samples.

### Quantitation of specific bacterial species using qPCR

To quantitatively validate the microbial community shifts, Syber Green qPCRs were performed using 2 μl of bacterial genomic DNA, in a total of 20 μl of reaction, from a subset of samples (n = 10) using an ABI Step One Plus Real time qPCR system software v2.3 (Applied-Biosystems). Primers were designed to determine the total bacterial concentration per sample per visit for *L. crispatus* (F’: 5’- GAT TTA CTT CGG TAA TGA CGT TAG GA −3’; R’: 5’-AGC TGA TCA TGC GAT CTG CTT TC –3)*, L. iners* (F’: 5’- TTG AAG ATC GGA GTG CTTGC −3’; R’: 5’- TTA TCC CGA TCT CTT GGG CA −3’)*, Prevotella bivia* (F’: 5’- CTG CGC TTG GTC CTG GTG GTT −3’; R’: 5’- CTT CAG GCG ACT CAA TAG GAC ACA AA −3’), and for pan-*Lactobacillus* spp. (F’: 5’- GTC CAT TGT GGA AGA TTC CC −3’; R’: 5’- TGG AAA CAG RTG CTA ATA CCG −3’), for total bacteria (16S region: F’: 5’- AAA CTC AAA KGA ATT GAC GG −3’; R’ 5’- CTC ACR RCA CGA GCT GAC −3’). Total concentrations of bacteria were inferred from standard curves as previously described^36^. Concentrations were reported as copy number/μl.

### Cytokine measurements using Luminex

The concentrations of 48 analytes were measured from CVLs using the Human Cytokine Panel A 48-Plex Discovery Assay® Luminex kit (Bio-Rad) according to the manufacturer’s instructions: Eotaxin, Fractalkine, GROα, IFNα2, IL-8, IP-10, MCP-1, MDC, MIG/CXCL9, MIP-1α, MIP-1β, RANTES, IL-1α, IL-1β, IL-6, IL-12p40, IL-12p70, IL-18, sCD40L, TNFα, TNFβ, MCP-3, IFN-γ, IL-7, IL-15, IL-9, IL-2, IL-4, IL-5, IL-17A, IL-17E/IL-25, IL-17F, IL-13, EGF, FGF-2, FLT-3L, G-CSF, GM-CSF, IL-3, M-CSF, PDGF-AA, PDGF-AB/BB, TGFα, VEGF-A, IL-1RA, IL-10, IL-22, and IL-27. A 5-parameter logistic regression was used to determine analyte concentrations from standard curves. If the concentration of a cytokine was lower than detectable level, it was given the value of half the lower than detectable value. If the analyte concentration was above the detectable level, it was given the value of the highest detectable concentration or the average of the highest detectable concentration and the highest standard dilution. The different analytes were categorized according to their primary biological function (Table 1).

**Table 1:** List of measured cytokines categorized as inflammatory, chemotactic, growth factors, adaptive cytokines, and regulatory cytokines.

| Function | Cytokines * |
| --- | --- |
| Inflammatory Cytokines | IFNγ, IL-1α, IL-1β, IL-6, IL-9, IL-12p40, IL-12p70, IL-18, MCP-3 (CCL7), sCD40 L, TNFα, TNFβ |
| Chemokines | Eotaxin (CCL11), Fractalkine (CX3CL1), GROα (CXCL1), IFNα2, IL-8 (CXCL8), IP-10 (CXCL10), MCP-1 (CCL2), MDC (CCL22), MIG/CXCL9, MIP-1α (CCL3), MIP-1β (CCL4), RANTES (CCL5) |
| Growth Factors | EGF, FGF-2, FLT-3L, G-CSF, GM-CSF, IL-3, M-CSF, PDGF-AA, PDGF-AB/BB, TGFα, VEGF-A |
| Adaptive Cytokines | IL-2, IL-4, IL-5, IL-7, IL-13, IL-15, IL-17A, IL-17E/IL-25, IL-17F |
| Regulatory Cytokines | IL-1RA, IL-10, IL-22, IL-27 |

\* Abbreviations: interferon (IFN); interleukin (IL); monochemoattractant protein (MCP); chemokine C-C motif ligand (CCL); soluble cluster of differentiation (sCD); tumour necrosis factor (TNF); chemokine ligand (CXCL); growth regulated alpha protein (GROα ); interferon gamma-induced protein (IP); macrophage derived chemokine (MDC); monokine induced by gamma interferon (MIG); macrophage inflammatory protein (MIP); regulated on activation, normal T-cell expressed and secreted (RANTES); epidermal growth factor (EGF); fibroblast growth factor (FGF); Fms-like tyrosine kinase 3 ligand (FLT-3L); granulocyte colony-stimulating factor (G-CSF); granulocyte-macrophage colony-stimulating factor (GM-CSF); macrophage colony-stimulating factor (M-CSF); platelet-derived growth factor (PDGF); transforming growth factor (TGF); and vascular endothelial growth factor (VEGF).

### Bioinformatics and statistical data analysis

Microbiota data analysis was performed using base R (v4.2.1) and relevant packages including phyloseq v1.40.0^37^, tidyverse^38^, vegan, and NMF. Within-participant alpha diversity was assessed using Chao1 index. To assess group-level differences in microbial composition, permutational multivariate analysis of variance analysis (PERMANOVA) was conducted on Bray-Curtis dissimilarities using the adonis function in the vegan package v2.6-2^39^. Pearson correlations were calculated using the stats package and visualized with ggpubr v0.4.0. Bonferroni correction was used for multiple comparisons.

All microbiome samples were categorized as ‘Optimal’ and ‘Non-optimal’ based on the sample’s *Lactobacillus* spp. dominance, microbial composition and diversity. In sub-analyses. longitudinal shifts in microbial composition were categorized as Positive, Negative or No change based on transitions in their *Lactobacillus* spp. abundance and their overall microbial composition and diversity. Two individuals with a ‘mixed’ shift (reduced overall diversity without a corresponding increase in *Lactobacillus* dominance, CST transition, or probiotic strain detection) and over 75% relative abundance of *Streptococcus anginosus* were excluded.

We were not statistically powered for analyses by treatment arm due to the small sample sizes within a Phase I study. Therefore, in some longitudinal analyses, participants were pooled across treatment arms and changes from baseline (Visit 2) to the end-of-intervention (Visit 4) were assessed for the entire cohort. In the absence of a non-intervention control arm, we applied a delta-based regression approach, calculating the per-participant change (Δ) in each outcome between end-of-intervention and baseline, allowing each participant to serve as their own control.

Multivariate logistic regression models, fitted using Δ-analytes as independent variables, were used to identify biomarkers associated with having a non-optimal vaginal microbiota. Eight cytokines/chemokines were positively associated with having a non-optimal microbiome: IL-1α, IL-1β, IL-6, IL-8, IL-18, MIP-1β, M-CSF, and TNFα. These eight biomarkers were used for construction of a composite inflammation score. The 75^th^ percentile value was calculated for each of these markers. For each participant, a value of 1 was assigned if the concentration of each individual analyte exceeded its respective 75th percentile, and 0 otherwise. The inflammation score was then calculated as the sum of these binary indicators, resulting in a discrete score ranging from 0 (no elevated analytes) to 8 (all analytes elevated).

The relationship between microbial taxa and genital cytokine concentrations were assessed using non-parametric Spearman’s rank correlations (ρ) between the relative abundances of the top bacterial taxa and concentrations of all measured analytes, due to its robustness to non-normal (right-skewed) distributions and ability to detect nonlinear monotonic relationships. The resulting correlation matrix was transformed into a distance matrix and subjected to hierarchical clustering with average linkage to identify groups of taxa exhibiting similar correlation patterns, hereafter referred to as Taxa Clusters (TCs). Clusters were defined using the hclust and cutreeDynamic functions, which balance sensitivity with stability in cluster detection. Significance of taxa–analyte correlations was determined using Benjamini corrections. Overall, p values <0.05 were considered statistically significant.

## Results

### Participant demographics and clinical characteristics

Genital microbiome data was available for 41/50 randomized participants (Table 2; Estring only: n = 9; Estring + vaginal RepHresh Pro-B: n = 9; Estring + oral RepHresh Pro-B: n = 10; vaginal RepHresh Pro-B: n = 13). All participants were of African, Caribbean, or Black ethnicity, with a median age of 35 years (interquartile range [IQR] 27 - 40). At baseline, 22.0% (n=9) were BV-positive and 17.1% (n=7) reported previously having had an STI, with none of them experiencing vaginal discharge. Overall, socio-behavioural, demographic, and clinical characteristics (sexual history, presence of PSA, relationship status, education) were similar across all study arms, suggesting successful randomization (Table 2).

**Table 2.** Characteristics of participants randomized to the four study arms using combinations of Estring and RepHresh Pro-B.

|  | **Overall (N = 41)** | **Estring (n = 9)** | **Estring + Oral Probiotic (n = 9)** | **Estring + Vaginal Probiotic (n = 10)** | **Vaginal Probiotic (n = 13)** | **P value** |
| --- | --- | --- | --- | --- | --- | --- |
| Age, median [IQR] | 35 [27 - 40] | 35 [31 - 37] | 31 [27 - 33] | 34 [25.5 - 39.75] | 38 [29 - 41] | 0.312 |
| ACB Ethnicity, n(%) | 41 (100) | 9 (100) | 9 (100) | 10 (100) | 13 (100) | - |
| Number of sex partners in last 6mo, n(%) |  |  |  |  |  | 0.530 |
| 0 | 16 (39.0) | 5 (55.6) | 3 (33.3) | 3 (30) | 5 (38.5) |  |
| 1-5 | 24 (58.5) | 3 (33.3) | 6 (66.7) | 7 (70) | 8 (61.5) |  |
| Choose not to respond | 1 (2.4) | 1 (11.1) | 0 (0) | 0 (0) | 0 (0) |  |
| Bacterial vaginosis at baseline | 9 (22.0) |  | 4 (44.4) | 1 (10) | 4 (30.8) | 0.090 |
| Previous history of bacterial vaginosis (Yes/No) | 10 (24.4) | 3 (33.3) | 0 (0) | 3 (30) | 4 (30.8) | 0.272 |
| 1 diagnosis | 5 (12.2) | 1 (11.1) | 0 (0) | 1 (10) | 3 (23.1) | 0.518 |
| >1 diagnosis | 5 (12.2) | 2 (22.2) | 0 (0) | 2 (15.4) | 1 (7.69) |  |
| Ever had an STI before study | 7 (17.1) | 1 (11.1) | 3 (33.3) | 2 (20) | 1 (7.69) | 0.493 |
| Irregular menstruation, n(%) | 38 (92.7) | 0 (0) | 0 (0) | 1 (10) | 2 (15.4) | 0.606 |
| Ever been pregnant, n(%) | 23 (56.1) | 4 (44.4) | 5 (55.6) | 5 (50) | 9 (69.2) | 0.685 |
| Condom usage, n(%) |  |  |  |  |  | 0.960 |
| All of the time (100% of the time) | 4 (9.8) | 0 (0) | 1 (11.1) | 2 (20) | 1 (7.69) |  |
| Almost every time (75-99% of the time) | 5 (12.2) | 1 (11.1) | 2 (22.2) | 0 (0) | 2 (15.4) |  |
| Most of the time (50-74% of the time) | 4 (9.8) | 1 (11.1) | 1 (11.1) | 1 (10) | 1 (7.69) |  |
| Sometimes (25-49% of the time) | 2 (4.9) | 1 (11.1) | 0 (0) | 0 (0) | 1 (7.69) |  |
| Rarely (less than 25% of the time) | 3 (7.3) | 1 (11.1) | 1 (11.1) | 1 (10) | 0 (0) |  |
| Never (0%) | 23 (56.1) | 5 (55.6) | 4 (44.4) | 6 (60) | 8 (61.5) |  |
| Presence of prostate specific antigen, n(%) |  |  |  |  |  |  |
| Baseline | 7 (17.1) | 2 (22.2) | 0 (0) | 2 (20) | 3 (23.1) | 0.534 |
| Mid-intervention | 7(17.1) | 1 (11.1) | 1 (11.1) | 1 (10) | 4 (30.8) | 0.597 |
| End of intervention | 4 (9.8) | 1 (11.1) | 0 (0) | 1 (10) | 2 (15.4) | 0.896 |
| Relationship status, n(%) |  |  |  |  |  |  |
| Married/Common law/Steady partner | 18 (43.9) | 2 (22.2) | 5 (55.6) | 5 (50) | 6 (46.2) | 0.678 |
| Single | 19 (46.3) | 5 (55.6) | 4 (44.4) | 5 (50) | 5 (38.5) |  |
| Divorced | 3 (7.3) | 2 (22.2) | 0 (0) | 0 (0) | 1 (7.69) |  |
| Widowed | 1 (2.4) | 0 (0) | 0 (0) | 0 (0) | 1 (7.69) |  |
| Highest level of education,<br>n(%) |  |  |  |  |  | 0.340 |
| Completed college or<br>university | 26 (63.4) | 5 (55.6) | 6 (66.7) | 6 (60) | 9 (69.2) |  |
| Completed secondary/high<br>school | 11 (26.8) | 4 (44.4) | 3 (33.3) | 3 (30) | 1 (7.69) |  |
| Completed elementary/primary school | 4 (9.8) | 0 (0) | 0 (0) | 1 (10) | 3 (23.1) |  |

### Genital microbial composition was mostly dominated by different *Lactobacillus* species

For a broad overview of the microbial composition among all participants at all visits, partitioning around medoids (PAM) was used to categorize all samples into four genital Community State Types (CST; Figure 2A). Overall, the CST III PAM cluster had a higher proportion of samples (58/14; 39.2%), followed by CST I and CST IV (both 29.1%). At treatment initiation (baseline), the largest proportion of samples (17/41; 41.5%) were categorized as CST III (*L. iners* dominant), 11/41 (26.8%) of samples were dominated by *L. crispatus* (CST I), while 12/41 (29.3%) had a microbiome dominated by BV-associated anaerobes (CST IV). Only one individual had a CST V (dominated by *L. jensenii*) microbiota (1/41 (2.44%)).

**Fig 2.**
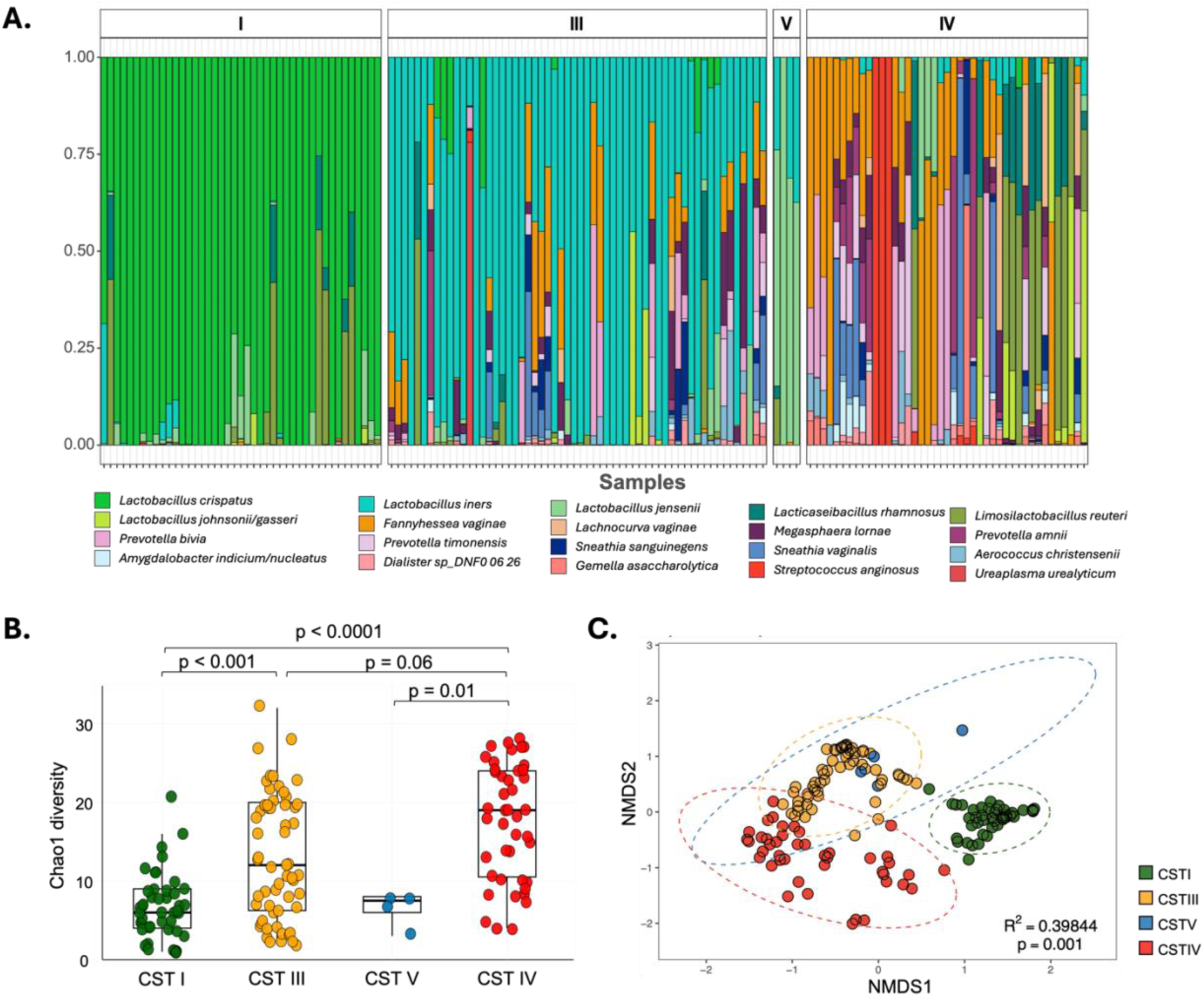
Differences in genital microbial community across community state types (CSTs) using partition around medoids for all participants and visits. **A.** Relative abundances of the 20 most abundant taxa by CST. **B.** Differences in alpha diversity/ within participant diversity (Chao1) among CSTs. **C.** Differences in beta diversity/between participant diversity (Bray-Curtis distances) by CST. CST I: Lactobacillus crispatus-dominant; CST III: L. iners-dominant; CST V: L. jensenii-dominant; CST IV: Diverse, mostly composed of BV-associated anaerobes.

In line with literature, CST I had the lowest alpha diversity (Figure 2B), followed by CST V. CST III (CST I vs CST III adj p = 0.004) and CST IV (CST I vs CST IV adj p < 0.0001; CST V vs CST IV adj p = 0.01; CST III vs CST IV adj p = 0.006) were associated with higher microbial diversity, statistically significantly after adjusting for multiple comparisons. Similarly, there was a distinct clustering in beta diversity among CSTs using Bray-Curtis distances (p = 0.001; Figure 2C).

### Both probiotic strains were detected in the genital tract with vaginally inserted, but not orally delivered, probiotics

To assess whether treatment with probiotics successfully colonized the vaginal tract, we compared the relative abundances of *L. rhamnosus* and *L. reuteri* from baseline to the final follow-up visit. Both probiotic taxa were confirmed to be absent across all study arms pre-treatment (Figure 3). Among participants randomized to the Estring-only or Estring + Oral probiotic arms, probiotic levels remained absent or below 2% throughout the study. In contrast, there was a significant increase in relative abundances within 15 days of treatment, by the mid-intervention visit, for the two vaginal probiotic arms: Estring + Vaginal probiotic (*L. rhamnosus*: p = 0.01; *L. reuteri*: p = 0.03; both: p = 0.03) and Vaginal probiotic only (*L. rhamnosus*: p < 0.01; *L. reuteri*: p < 0.01; both: p < 0.01). Further, for the Vaginal probiotic-only arm, median relative abundances remained elevated till the end-of-intervention visit (*L. rhamnosus*: p = 0.02; *L. reuteri*: p = 0.05; both: p = 0.03). However, following product discontinuation, probiotic lactobacilli levels returned to baseline, falling below 2% or becoming undetectable on both vaginal probiotic arms (Figure 3).

**Fig 3.**
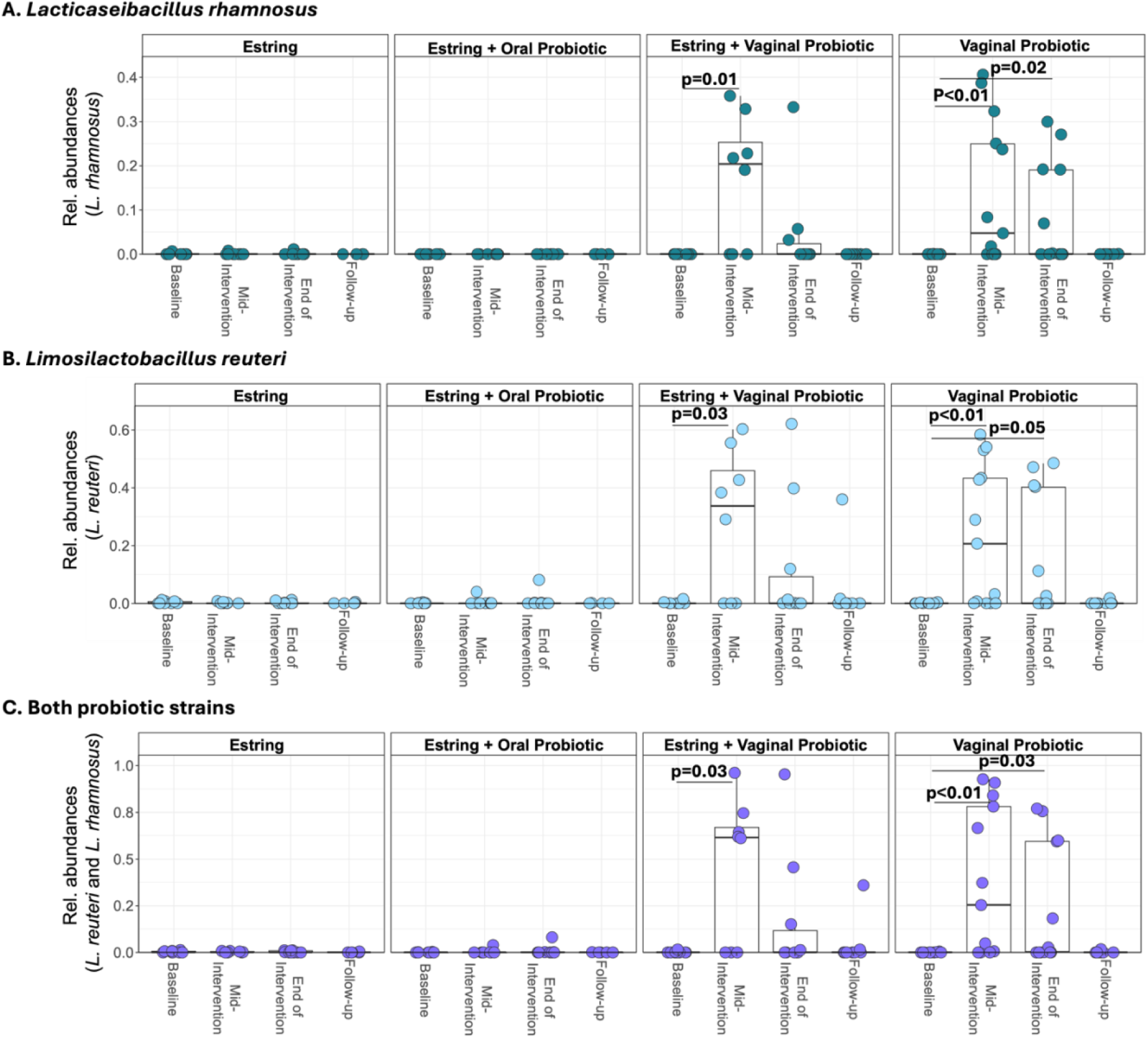
Relative abundances of the probiotic species **A.** Lacticaseibacillus rhamnosus, **B.** Limosilactobacillus reuteri, or **C.** the cumulative abundance of both taxa, at all study visits for the different treatment arms. Mann-Whitney U-tests were used to compare the relative abundances to baseline. P values <0.05 were considered statistically significant.

### Longitudinal increases in *Lactobacillus* spp. and shifts toward lower diversity among all participants

In addition to the increase seen in the relative abundances of the probiotic taxa, we found an increase in overall *Lactobacillus* abundances from baseline (median 60.9%; Figure 4A) to the mid- and end-of-intervention visits (72.3% and 70.5% respectively). In line with this, there was an increase in the number of participants with an overall *Lactobacillus-*high microbiota over time (Baseline: 61.0%; mid-intervention: 64.1%; end-of-intervention: 70.7%; Figure 4B). More importantly, despite the loss of the probiotic lactobacilli post-treatment, overall abundances of all *Lactobacillus* taxa remained higher at the follow-up visit, a week after treatment ended (68.9%), compared to baseline. This was accompanied by a decrease in microbial diversity (p = 0.044; Figure 4C). Among participants with samples at all four visits (N = 25), there was an increase in the proportion of the *L. crispatus-*dominant CSTs at the end-of-intervention and at the follow-up visit compared to the baseline visit (Baseline: 24%; end-of-intervention: 28% and follow-up visit: 32%; Figure 4D) as well as a decrease in the more diverse CST IV (Baseline: 28%; end-of-intervention: 28% and follow-up visit: 24%).

**Fig 4.**
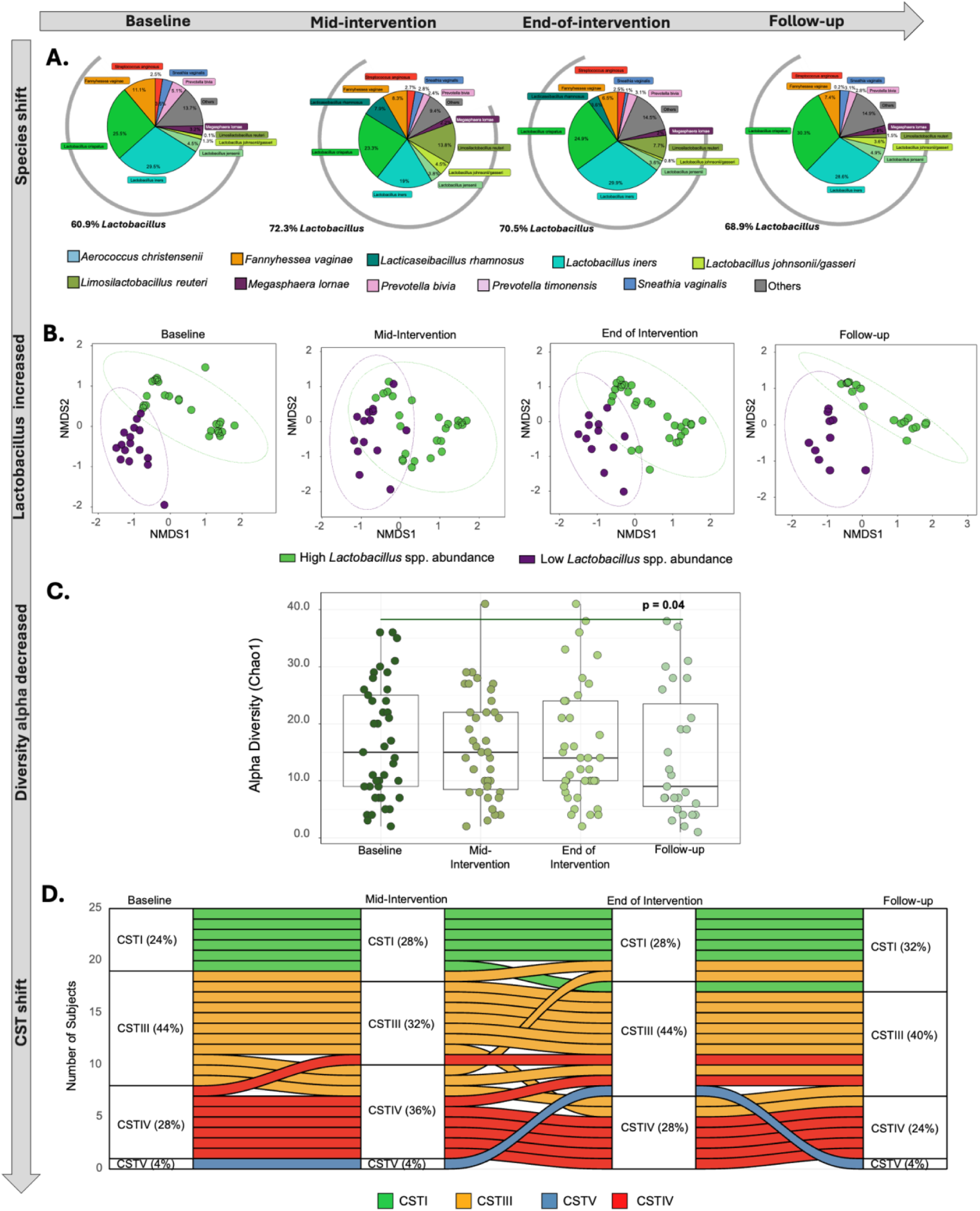
Longitudinal changes in microbiome composition across study visits for all participants. **A.** Comparing the overall microbial composition from baseline to the follow-up visit, particularly for Lactobacillus spp. **B.** Bray-Curtis distances showing that samples cluster according to Lactobacillus spp. abundance, with an increasing number of samples within the Lactobacillus-high cluster. PERMANOVA was used to test for significant differences between the clusters. **C.** Change in Chao1 alpha diversity across all visits. Mann-Whitney U-tests were used to compare alpha diversities at different visits to the baseline. **D.** Alluvial plot showing CST shifts from baseline to the follow-up visit. For all statistical tests, p < 0.05 was considered statistically significant.

### Shifts towards a more optimal microbiota in majority of participants following intervention

Next, to assess the longitudinal effectiveness of the different interventions in improving microbial composition, baseline and end-of-intervention samples were categorized as ‘Optimal’/’Non-optimal’ based on the sample’s CST, alpha diversity, and dominance of any *Lactobacillus* spp. Overall, a higher percentage of samples had an ‘Optimal’ microbial composition (28/41; 68.3%) at the end-of-intervention visit compared to 20/41 (48.7%) at the baseline visit (Figure S1).

Further, to measure these shifts at an individual level, participants were categorized as either having an overall positive change, a negative change, or no change, based on four criteria in this particular order: (1) A transition to a less diverse CST (5/41); (2) A shift to a higher *Lactobacillus* spp. relative abundance quartile (16/41); (3) A decrease in overall alpha diversity based on bacterial taxa other than Lactobacillus spp. (20/41); and (4) A decrease in BV-associated bacteria, indicating a transition towards a less dysbiotic vaginal environment (16/41) – in cases where few BV-associated bacteria dominated at baseline (leading to a low alpha diversity; Figure 5A) Most participants showed overlap across the different criteria, with two individuals fulfilling all four criteria (Figure 5B).

**Fig 5.**
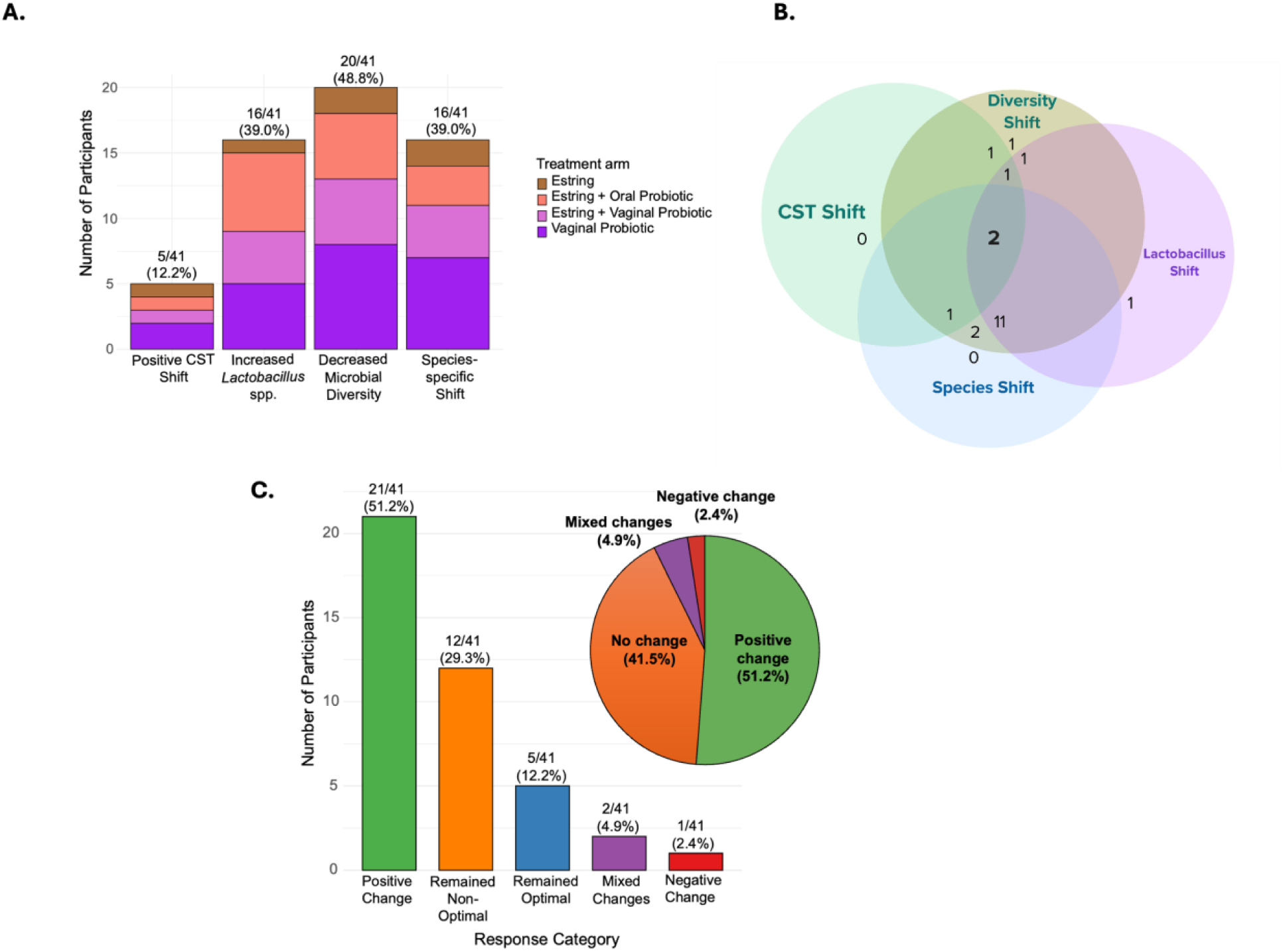
Defining overall changes in the genital microbiota from baseline to end-of-intervention. **A.** Frequency of participants fulfilling each of the four criteria used to define longitudinal improvement in the genital microbiota with treatment. **B.** Venn diagram showing the overlap of these criteria across samples. **C.** Number of participants categorized as experiencing a positive change/improvement, no change (remaining optimal or non-optimal), having a mixed profile, or having a negative change over the 30 days of treatment.

By the end-of-intervention visit, a larger proportion of participants across all study arms exhibited a general positive transition in their microbiome composition (21/41 (51.25%); Figure 5C), while 19 individuals (46.3%) showed no change in their vaginal microbiota from the first visit. One participant from the Estring + vaginal probiotic arm experienced a negative shift, where they transitioned from *L. crispatus* (CST I; relative abundances of *L. crispatus* = 68.7% and *L. iners* = 30.9%), to *L. iners* dominance (CST III; relative abundances of *L. crispatus* = 33.7% and *L. iners* = 64.3%). Notably, they did return to CST I post-treatment (relative abundances of *L. crispatus* = 94.0% and *L. iners* = 0.4%). Further, two participants (from the two vaginal probiotic arms) experienced a ‘mixed’ shift defined by a decrease in total diversity without a corresponding increase in probiotic strain detection, CST transition, or *Lactobacillus* dominance.

Dabee et al. **Fig S1.** Comparing the relative abundances of the 20 most abundant taxa between samples categorized as Non-optimal and Optimal at the **A.** Baseline and **B.** End-of-intervention visits. Sample sizes and percentages of samples categorized as Optimal/Non-optimal are reported.

### Participants who responded positively had higher abundances of *Lactobacillus* spp. and lower abundances of BV-associated bacteria

Among participants categorized as having a longitudinal positive shift as described above, as expected, we found a larger increase in overall *Lactobacillus* abundances from baseline (median 29.3%; Figure 6A) to the mid- and end-of-intervention visits (71.5% and 87.8% respectively). Crucially, at the follow-up visit, 7 days after treatment stoppage, this increase in lactobacilli was sustained (63.4% *Lactobacillus* spp. relative abundance), primarily due to an increase in the relative abundance of *L. iners.* This was accompanied by a decrease in the BV associated anaerobes found at higher abundances at baseline, such as *Fannyhessea vaginae* (21.4% to 2.4%)*, Prevotella bivia* (10.8% to 1.6%)*, Megasphaera lornae* (9.6% to 9.1%)*, P. timonensis* (7.4% to 2.1%) and *Sneathia vaginalis* (3.2% to 0.7%), indicating that once treatment was stopped, depletion of the probiotic taxa was replaced by other lactobacilli rather than pathogenic anaerobes up to a week later, the final time point of follow up. This was reflected in Figure 6B, with clustering of larger numbers of *Lactobacillus*-high individuals both at the end-of-intervention and follow-up visits compared to baseline. In line with the increases seen in *Lactobacillus* spp. abundances, participants with an improved genital microbiome exhibited a statistically significantly lower alpha diversity (p = 0.03; Figure 6C) and CST transitions away from the diverse, BV-associated CST IV to *Lactobacillus* dominance (CST I, III, or V; Figure 6D).

**Fig 6.**
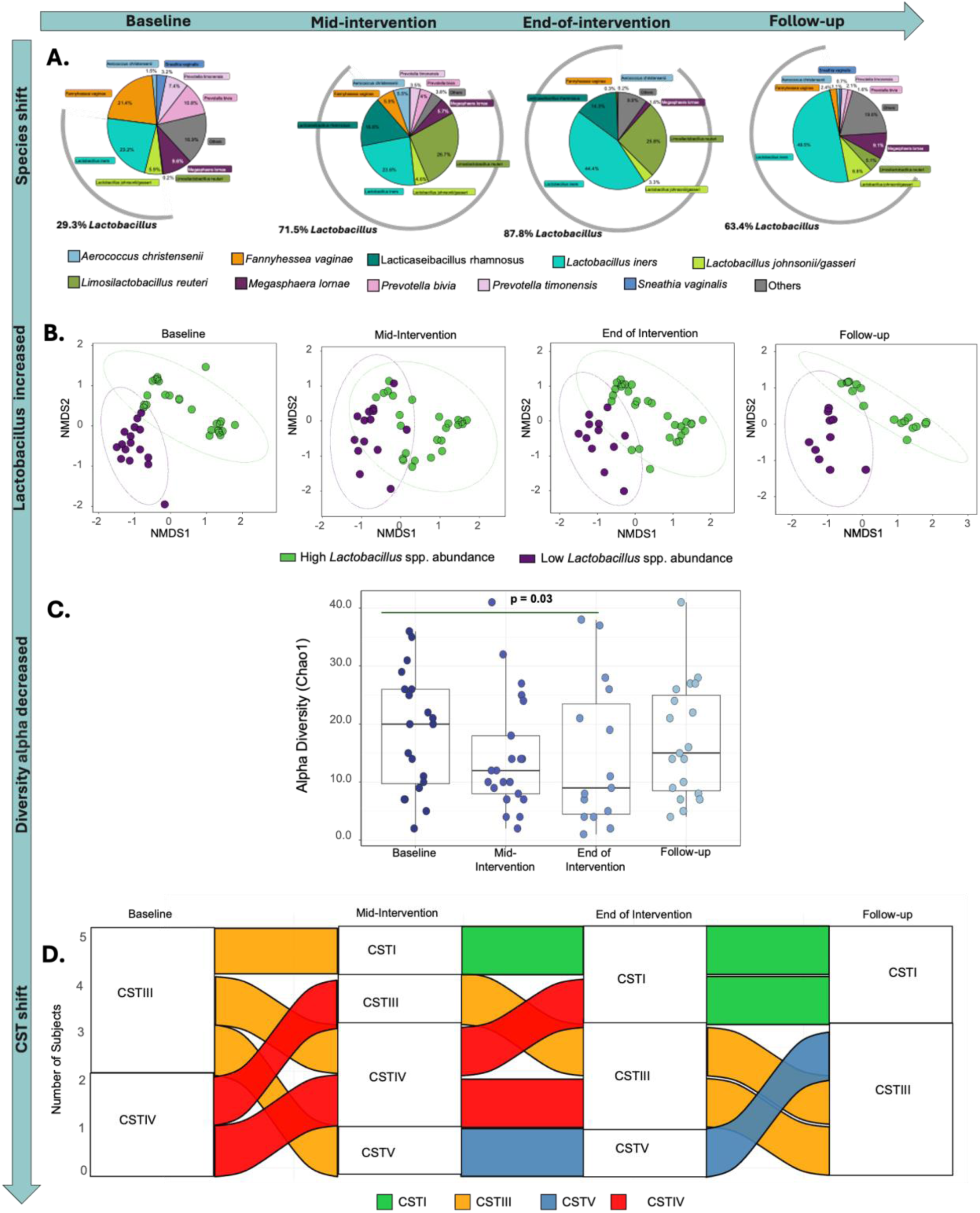
Longitudinal changes in microbiome composition across study visits for participants who experienced a longitudinal improvement in microbial composition. **A.** Comparing the overall microbial composition from baseline to the follow-up visit, particularly for Lactobacillus spp. **B.**

To confirm that this increase in *Lactobacillus* spp. abundance and decreases in BV-associated bacteria were not simply due to the compositional nature of sequencing data, quantitative qPCRs were carried out to determine the concentrations of lactobacilli and the BV-associated *P. bivia*. Among ten randomly selected participants who exhibited a positive shift from baseline to the end of the intervention (Figure S2), the total concentrations of *L. crispatus*, as well as all lactobacilli appeared to increase, while the concentration of P. bivia tended to decrease, in line with the 16S rRNA gene sequencing findings. However, these differences were not statistically significant, likely due to the small sample size. Further, we found no difference in the total bacterial load from baseline to post-treatment, suggesting a shift in overall bacterial composition rather than an overgrowth of individual bacteria (Figure S2E).

Bray-Curtis distances showing that samples cluster according to Lactobacillus spp. abundance, with an increasing number of samples within the Lactobacillus-high cluster. PERMANOVA was used to test for significant differences between the clusters. **C.** Change in Chao1 alpha diversity across all visits. Mann-Whitney U-tests were used to compare alpha diversities at different visits to the baseline. **D.** Alluvial plot showing CST shifts from baseline to the follow-up visit. For all statistical tests, p < 0.05 was considered statistically significant.

Dabee et al. **Fig S2.** Validation of key taxonomic shifts using bacterial-specific qPCR assays. Changes in the total bacterial concentration of **A.** All Lactobacillus spp., **B.** Lactobacillus crispatus, **C.** Lactobacillus iners, and **D.** Prevotella bivia were quantified by qPCR. Paired t-tests were performed on a random subset (n=10) of participants who experienced a positive microbial transition from baseline to the end of the intervention, confirming the trends observed in sequencing data. A p-value of <0.05 was considered statistically significant.

### Overall, treatment was associated with reduced inflammation, particularly among those with an improved microbiome composition

Forty-eight different cytokines and soluble immune markers concentrations were measured across all visits for all participants. Longitudinal differences in the concentrations of each of the 48 different soluble immune markers measured in this study are shown in Figure 7A. Using a multivariate logistic regression, we found that 13 of these markers were statistically significantly associated with having a non-optimal microbiome. Five cytokines - IL-1RA, GROα, G-CSF, MIG, and IP-10 were negatively associated with having a non-optimal microbiota (as shown by the green asterisks in Fig 7A), while eight markers of inflammation were positively associated with having a non-optimal microbiome: IL-1α, IL-1β, IL-6, IL-8, IL-18, MIP-1β, M-CSF, and TNFα (red asterisks). Longitudinal changes in the inflammation score were determined based on the concentrations of these 8 cytokines/chemokines, as described in Materials and Methods.

**Fig 7.**
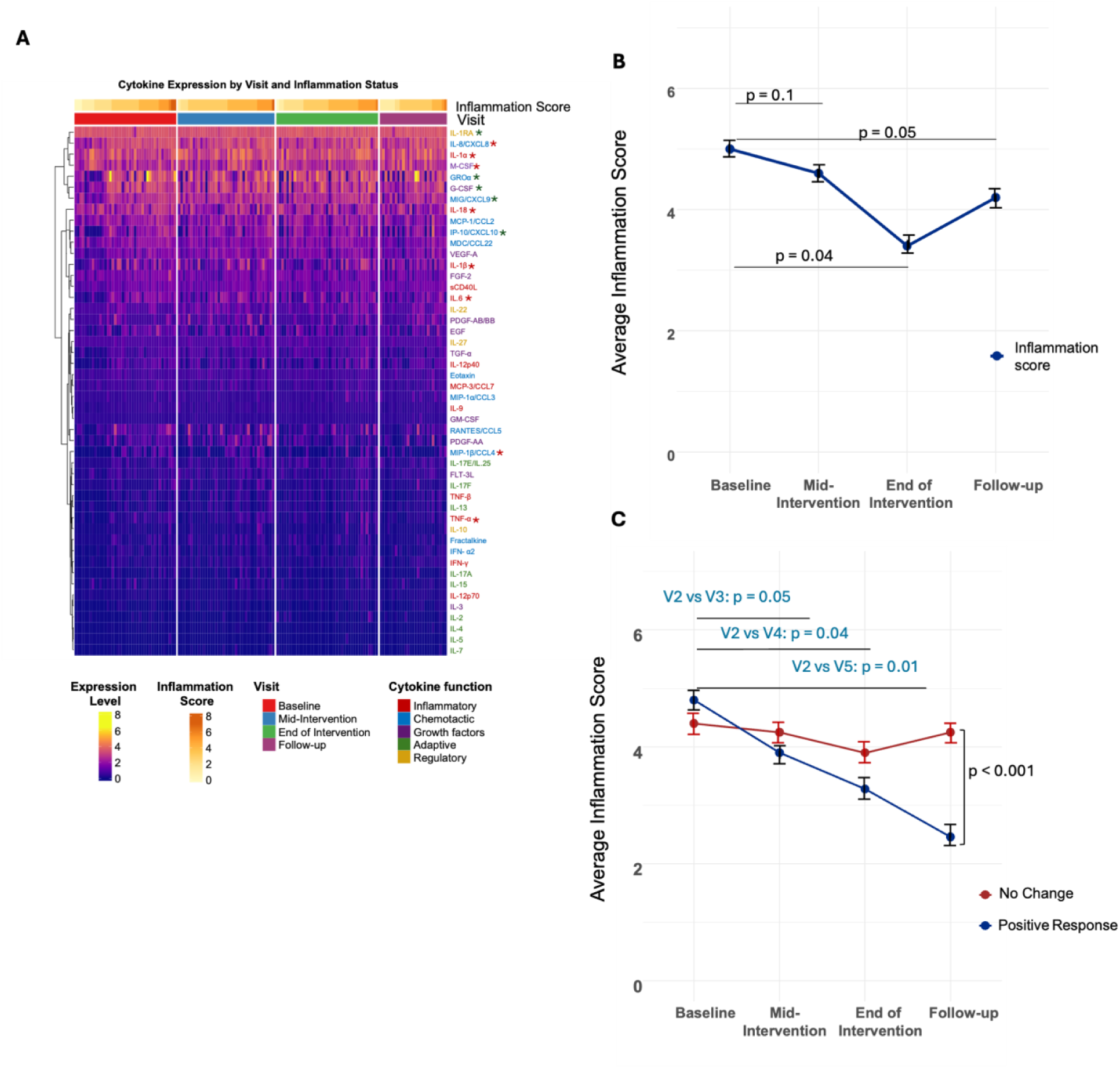
Change in inflammation score across visits with treatment. **A.** Cytokine heatmap showing overall concentration changes from baseline to end-of-intervention and at the follow-up visit. Red asterisks indicate a significant positive association with the non-optimal group, while green asterisks indicate a significant negative association with the non-optimal group (and positive association with the optimal group) (p < 0.05). Analytes positively associated with a non-optimal microbiome (red asterisks) were used to generate a composite inflammatory score. **B.** Overall change in average inflammation score from baseline to follow-up for all participants. **C.** Difference in inflammation score at each visit between participants who had a positive change and those who did not experience a change in microbiome composition. Mann-Whitney U-tests were used for comparisons. P < 0.05 was considered statistically significant.

There was a consistent decrease in overall inflammation from baseline (average score: 4.4) to mid-intervention (average score: 4.2) and end-of-intervention (average score: 3.8; p = 0.04; Figure 7B). Treatment stoppage was associated with an increase in inflammation, although it remained lower than at baseline (average score at follow-up: 4.1; p = 0.05). We observed a significant difference in the trajectory of inflammation scores between participants who exhibited a positive shift in their microbiota following treatment and those who experienced no change (Figure 7C). While there were no statistically significant shifts across visits for those without a positive change, participants with a positive change had a consistently lower score at every visit compared to baseline (average score: 4.8), with this trend continuing up to a week later post-treatment (mid-intervention average score: 3.9, p = 0.05; end-of-intervention average score: 3.28, p = 0.04; follow-up visit average score: 2.46, p = 0.01). Additionally, there was a marked difference in inflammation scores by the follow-up visit when comparing the positive response and no-change groups despite treatment stoppage (p < 0.001; Figure 7C).

### Specific cytokine signatures are associated with optimal and non-optimal microbiome communities

To carry out integrative analyses, four distinct bacterial taxon clusters (TC1 – TC4) were identified using hierarchical clustering of Spearman correlations between bacterial relative abundances (Figure 8A). TC1, TC2, and TC4, largely composed of anaerobic taxa classically associated with BV, were positively correlated with each other. These three clusters were all positively associated with proinflammatory cytokines (IL-1α, IL-1β, IL-8, IL-18, TNFα) and negatively associated with anti-inflammatory biomarkers (IL-1RA, IL-4, IP-10, MIG, MDC, GROα) (Figure 8B). TC3, in contrast, was exclusively composed of lactic acid-producing *Lactobacillus* species, consistent with a more optimal, protective vaginal microbiome: *L. crispatus, L. gasseri, L. jensenii, L. reuteri, L. coleohominis, L. rhamnosus,* and *L. johnsonii.* This *Lactobacillus* spp. cluster was associated with distinct cytokine correlation patterns (Figure 8B) comprised of lower concentrations of proinflammatory cytokines and higher levels of anti-inflammatory cytokines. Of note, *L. iners* was clustered within TC2, with taxa associated with BV genital inflammation, suggesting some shared ecological and immunological characteristics with a more diverse microbiota.

**Fig 8.**
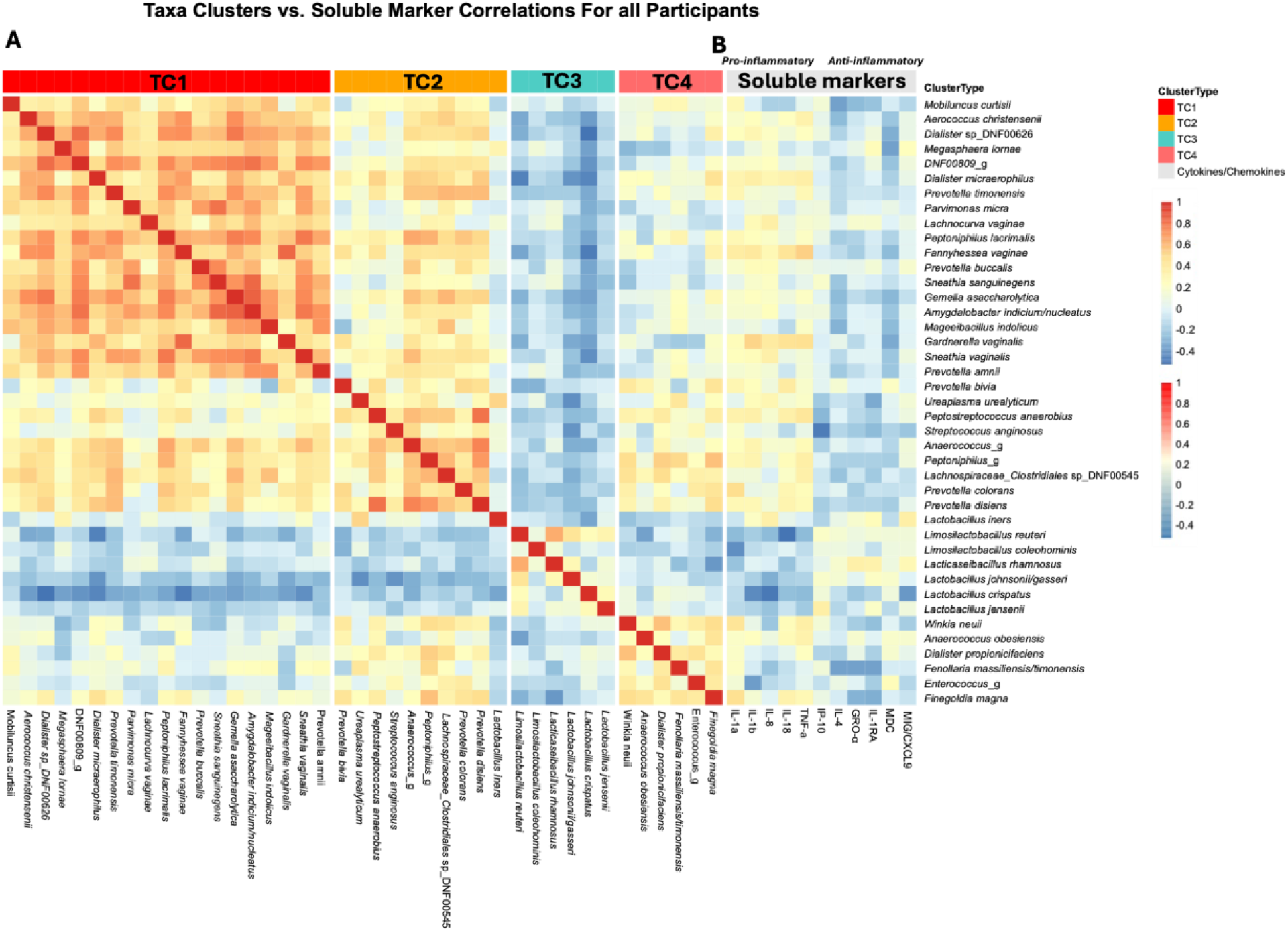
Correlations between all bacterial taxa and soluble mediators associated with inflammation. **A.** Four clusters of positively associated bacterial taxa (TC1 – TC4) were identified using Spearman correlations **B.** Spearman correlations between bacterial taxa and cytokines associated with inflammation (either Pro-inflammatory or Anti-inflammatory)

## Discussion

In this randomized, open-label, Phase I clinical trial, we evaluated the impact of low-dose intravaginal estrogen, combined with different probiotic delivery routes of *L. rhamnosus* GR-1 and *L. reuteri* RC-14, on the genital microbiome and immune milieu of premenopausal women. While other studies have examined the safety and effectiveness of probiotic strains previously^40–42^, this study is the first to directly compare these delivery routes in combination with estrogen and show the safety and feasibility^29^. Here, we report the changes in biological outcomes based on the treatments. While there was an overall shift towards a less diverse microbiome among most participants across treatment arms, we demonstrate that vaginal, but not oral, administration of probiotics led to consistent detection of these taxa in the genital tract, significant change in microbial composition, and reduced inflammation. Further, in our study, treatment led to lower inflammation levels, significantly so among those with an overall positive shift in microbial composition.

Several studies have explored the use of different *Lactobacillus* strains as vaginal or oral probiotics, mostly aimed at reducing BV incidence and recurrence. Women given vaginal *L. crispatus* CTV-05 (Lactin-V), post metronidazole, had a 34% lower risk of BV recurrence 12 weeks later, and the Lactin-V probiotic strain could be detected in 79% of participants up to 3 months post treatment stoppage^26^. Similarly, the recent VIBRANT Phase I study, assessing the efficacy of a multi-strain *L. crispatus* vaginal tablet, also showed successful colonization in 66% of participants, with strains detectable up to 12 weeks post-treatment^43^. In our study, *L. rhamnosus* GR-1 and *L. reuteri* RC-14 were evaluated due to their demonstrated ability to colonize the vagina, exert a broad range of antimicrobial effects, and potentially improve vaginal health by inhibiting the growth of pathogens like Candida and Gardnerella^41,42,44,45^. Here, the ability of probiotic strains to colonize vaginally, but not orally, in our study supports prior findings^46–48^. However, the fact that colonization by probiotic strains was transient indicates that sustained clinical benefits may require ongoing or long-term supplementation regimens.

Although colonization by the probiotic strains diminished after treatment cessation, the broader dominance of endogenous *Lactobacillus* spp. and lower microbial diversity persisted by the last week of the study, one week post-treatment, supporting the protective role of probiotic lactobacilli, even if not *L. crispatus* specifically. This shift from probiotic to endogenous lactobacilli suggests that transient colonization may act as a “catalyst,” creating conditions favorable for lactobacilli colonization. Importantly, the intervention led to an enrichment of both the proportion, and the overall concentration of total lactobacilli, suggesting that the underlying mechanism may rely on the competitive exclusion of dysbiotic bacteria. This could occur through stronger adhesion by some *Lactobacillus* spp. or through the production of antimicrobial mediators, preventing the growth of BV-associated dysbiotic bacteria^49^. This compositional shift, even in the absence of antibiotics, is a promising ecological approach to restoring vaginal health without the microbial disruptions associated with antibiotic use. Interestingly, women using both the Estring and vaginal probiotic had lower abundances of probiotic taxa at end-of-intervention despite the overall increase in *Lactobacillus* spp. abundance, suggesting that estrogen may play a more nuanced role in modulating colonization by exogenous strains of lactobacilli in the genital tract. Crucitti et al. (2018) found an increase in *Lactobacillus*-dominance with the use of estrogen-containing vaginal rings^50^, which suggests that the combination of estrogen and vaginal probiotic may have influenced the shift to endogenous lactobacilli, compared to the vaginal probiotic only arm. These dynamics underscore the complex interplay between exogenous probiotic species, endogenous lactobacilli, and hormonal context in shaping long-term colonization patterns. However, there was some variability in the participants’ responses to the intervention. Notably, treatment was associated with a temporary negative shift in one participant, marked by reduced lactobacilli and increased BV-associated anaerobes. However, this disruption was rapidly reversed at follow-up, with a return to a *lactobacillus*-dominated state.

More importantly, we demonstrated that a targeted microbiome intervention can lead to clinically relevant immune improvements: transitioning to Lactobacillus-dominated CSTs was associated with consistent reductions in key proinflammatory cytokines, an effect that persisted even after treatment cessation. Given the well-established links between BV-associated dysbiotic microbiota, chronic cervicovaginal inflammation, adverse reproductive outcomes, and an increased risk of STI and HIV acquisition^6,51,52^, our findings provide further evidence that strategies that target microbiome shifts can improve both microbial dysbiosis and decrease host inflammation. This is in line with previous findings from the Lactin-V trial reporting a decrease in concentrations of IL-1α and soluble E-cadherin^53^. This dual effect was further demonstrated by our correlation analyses, that linked BV-associated anaerobes to a proinflammatory biomarker signature while identifying a cluster of *Lactobacillus* species associated with lower concentrations of these proinflammatory biomarkers and positively correlated to higher concentrations of anti-inflammatory markers. Unlike other lactobacilli, *L. iners* clustered with BV-associated taxa, consistent with its ambiguous role as a transitional species frequently detected in both optimal and dysbiotic states^54^.

This study had several limitations. The modest sample size, due to it being a phase I trial carried out during the COVID-19 lockdown (2020-2021), limited our statistical power to detect smaller effect sizes and assess differences across treatment subgroups. Another caveat was the lack of a no-intervention control arm to act as the reference group. Although we identified strong correlations between specific microbial communities and cytokine profiles, the directionality of these relationships still needs to be investigated further.

Despite the limitations, the results from the current study will directly inform the design of the next phase of the study. While the present study was not powered for definitive comparisons between intervention arms, direct vaginal probiotic delivery was superior to oral when targeting vaginal colonization. In future studies, comparing the effect of vaginal interventions on both individuals with and without BV may allow us to better assess the impact of probiotic lactobacilli on a broader range of diverse microbiome compositions. Further, including a control group will enable the comparison of longitudinal changes in microbiota across the different treatment arms to a no-intervention reference group. A well-powered Phase II trial will allow us to determine the optimal intervention strategy and its efficacy in improving vaginal health. These results will help assess whether these microbial and immunological changes translate into clinically relevant outcomes, such as reduced recurrence of BV, decreased inflammation, or lowered susceptibility to sexually transmitted infections.

In conclusion, this Phase I trial provides critical proof-of-concept evidence that the vaginal microbiome and immune milieu can be positively modulated through a targeted intervention, with the route of administration being an influential factor. The fact that a decrease in inflammation was observed in our study using a probiotic other than *L. crispatus* indicates that various *Lactobacillus* spp. are viable options for creating an environment conducive to subsequent colonization by endogenous lactobacilli. This strategy could offer a novel, non-antibiotic-dependent therapeutic method to address the significant global burden of BV and its associated health disparities.

## Data Availability

All data produced in the present study are available upon reasonable request to the authors

## Acknowledgements

The authors would like to acknowledge the technical help of Sussan Kianpour, Tushar Dhawan, Andrew Rempel in processing the CVL samples. We would also like to acknowledge the excellent help from the CTN team led by Judith Needham for their assistance and oversight of the trial, including data management plan, data safety management board, Health Canada protocol development, review and submission, clinical site monitoring visits, and monthly progress monitoring and risk management meetings. Analyses were enabled in part by support provided by Graham cloud (graham.computecanada.ca) and the Digital Research Alliance of Canada (alliancecan.ca).

## Author contributions

Conceptualisation: CK, FS, GR, WT

Data curation: SD, MN, BG, CH, JW

Formal analysis: SD, MN, BG, AN, JW, CH

Funding acquisition: CK, FS, WT

Investigation: CK, FS, RK, JW, WT

Methodology: CH, BG, JW,

Project administration: CK, FS, JR, MA, JR, WT, RK

Resources: CK, FS, WT

Software: SD, MN, BG, CH

Supervision: CK, FS, JR, WT

Validation: JR, CH, BG

Visualisation: SD, MN, BG

Writing – original draft: SD, MN, CK

Writing – review & editing: FS, GR, RK,

## Declaration of interest

We have no conflicts of interest to disclose.

## Data sharing

The sequencing reads generated during this study have been deposited in the NCBI Sequence Read Archive (SRA): BioProject ID PRJNA1372819. Associated de-identified data and data dictionary can be made available upon request.

## Role of funding source

This study was funded by CIHR Grants FRN#159229 and FRN#154047 (CK) and in-kind contributions from CIHR HIV Clinical Trials Network (FS, CK). There was no additional external funding received for this study. The funders had no role in study design, data collection and analysis, decision to publish, or preparation of the manuscript.

## Notes

### Competing Interest Statement

The authors have declared no competing interest.

### Clinical Trial

NCT03837015

### Author Declarations

Ethics approval was obtained from the Hamilton Integrated Research Ethics Board (HiREB Project #7061).

