## Supplementary Figures for "Changes in the Genital Microbiome and Inflammation Among Women Using 30-Day *L. Rhamnosus/L. Reuteri* Probiotic in Combination with Intravaginal Estradiol: Findings from a Pilot, Open-Label, Phase I Clinical Trial"

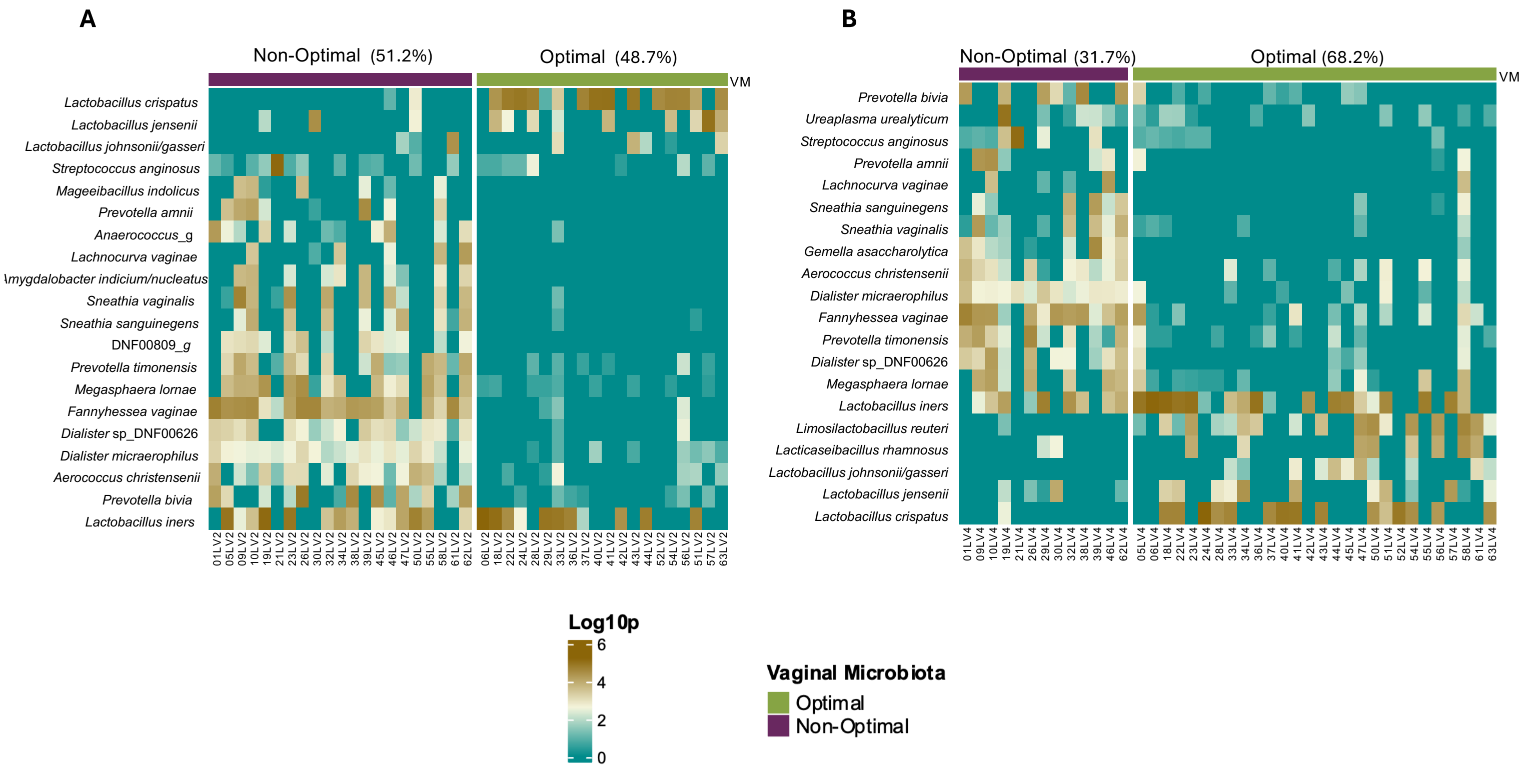


**Figure S1.** Comparing the relative abundances of the 20 most abundant taxa between samples categorized as Non-optimal and Optimal at the A) Baseline and B) End-of-intervention visits. Sample sizes and percentages of samples categorized as Optimal/Non-optimal are reported.

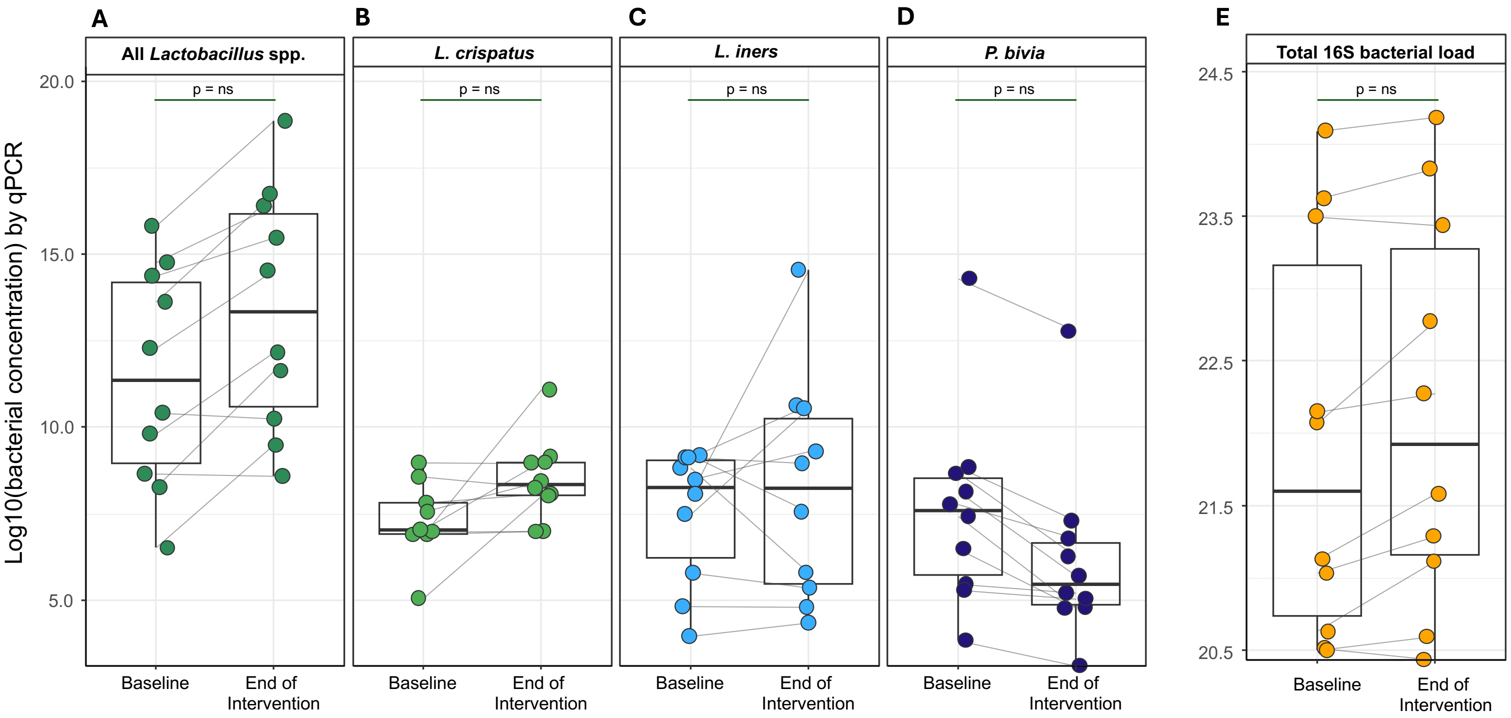


**Figure S2. V**alidation of key taxonomic shifts using bacterial-specific qPCR assays. Changes in the total bacterial concentration of (A) All *Lactobacillus* spp., (B) *Lactobacillus crispatus*, (C) *Lactobacillus iners*, and (D) *Prevotella bivia* were quantified by qPCR. Total 16S concentrations (D) were calculated as a proxy of total bacterial burden per sample. Paired t-tests were performed on a random subset (n=10) of participants who experienced a positive microbial transition from baseline to the end of the intervention, confirming the trends observed in sequencing data. A p-value of <0.05 was considered statistically significant.
